# Deep learning for AI-based diagnosis of skin-related neglected tropical diseases: a pilot study

**DOI:** 10.1101/2023.03.14.23287243

**Authors:** Rie Yotsu, Zhengming Ding, Jihun Hamm, Ronald Blanton

**Affiliations:** Department of Tropical Medicine, Tulane University School of Public Health and Tropical Medicine, 1440 Canal Street, New Orleans, LA 70112; Department of Computer Science, Tulane University School of Science and Engineering, 201 Lindy Claiborne Boggs Center, 6823 St. Charles Avenue, New Orleans, LA 70118

**Keywords:** artificial intelligence, Buruli ulcer, deep learning, Hansen’s disease, leprosy, machine learning, mycetoma, scabies, skin, skin NTDs, skin of color, yaws

## Abstract

**Background:** Deep learning, which is a part of a broader concept of artificial intelligence (AI) and/or machine learning has achieved remarkable success in vision tasks. While there is growing interest in the use of this technology in diagnostic support for skin-related neglected tropical diseases (skin NTDs), there have been limited studies in this area and fewer focused on dark skin. In this study, we aimed to develop deep learning based AI models with clinical images we collected for five skin NTDs, namely, Buruli ulcer, leprosy, mycetoma, scabies, and yaws, to understand how diagnostic accuracy can or cannot be improved using different models and training patterns.

**Methodology:** This study used photographs collected prospectively in Côte d’Ivoire and Ghana through our ongoing studies with use of digital health tools for clinical data documentation and for teledermatology. Our dataset included a total of 1,709 images from 506 patients. Two convolutional neural networks, ResNet-50 and VGG-16 models were adopted to examine the performance of different deep learning architectures and validate their feasibility in diagnosis of the targeted skin NTDs.

**Principal findings:** The two models were able to correctly predict over 70% of the diagnoses, and there was a consistent performance improvement with more training samples. The ResNet-50 model performed better than the VGG-16 model. Model trained with PCR confirmed cases of Buruli ulcer yielded 1-3% increase in prediction accuracy over training sets including unconfirmed cases.

**Conclusions:** Our approach was to have the deep learning model distinguish between multiple pathologies simultaneously – which is close to real-world practice. The more images used for training, the more accurate the diagnosis became. The percentages of correct diagnosis increased with PCR-positive cases of Buruli ulcer. This demonstrated that it may be better to input images from the more accurately diagnosed cases in the training models also for achieving better accuracy in the generated AI models. However, the increase was marginal which may be an indication that the accuracy of clinical diagnosis alone is reliable to an extent for Buruli ulcer. Diagnostic tests also have its flaws, and they are not always reliable. One hope for AI is that it will objectively resolve this gap between diagnostic tests and clinical diagnoses with addition of another tool. While there are still challenges to be overcome, there is a potential for AI to address the unmet needs where access to medical care is limited, like for those affected by skin NTDs.

**AUTHOR SUMMARY:** The diagnosis of skin diseases depends in large part, though not exclusively on visual inspection. The diagnosis and management of these diseases is thus particularly amenable to teledermatology approaches. The widespread availability of cell phone technology and electronic information transfer provides new potential for access to health care in low-income countries, yet there are limited efforts targeting these neglected populations with dark skin and consequently limited availability of tools. In this study, we leveraged a collection of skin images gathered through a system of teledermatology in the West African countries of Côte d’Ivoire and Ghana, and applied deep learning, a form of artificial intelligence (AI) - to see if deep learning models can distinguish between different diseases and support their diagnosis. Skin-related neglected tropical diseases, or skin NTDs, prevail in these regions and were our target conditions: Buruli ulcer, leprosy, mycetoma, scabies, and yaws. The accuracy of prediction depended on the number of images that were fed into the model for training with marginal improvement using laboratory confirmed cases in training. Using more images and greater efforts in this area, it is possible that AI can help address the unmet needs where access to medical care is limited.

## INTRODUCTION

Deep learning has achieved remarkable success in vision tasks such as image classification, image localization and image semantic segmentation, which also includes skin disease prediction. Deep learning is a part of a broader concept of artificial intelligence and/or machine learning whereby it uses vast volumes of data and complex algorithms to train a model to perform certain tasks. The success of the approach undoubtedly can be attributed to the ability of learning abstract semantic knowledge with the hierarchical network architecture from visual signals (1). It is increasingly gaining interest and becoming more important in the field of dermatology in this digital era. Evidence is accumulating that deep learning can assist healthcare providers to make better clinical decisions, even to an extent that sometimes exceeds human judgement (2, 3, 4). However, many of the diseases studied are pigmented lesions such as melanoma and basal cell carcinoma, or inflammatory dermatoses which often affect people with lighter skin color and thus provide a high degree of contrast (5, 6, 7).

Skin-related neglected tropical diseases, or skin NTDs, comprise a group of infectious diseases whose morbidity is expressed on the skin. They include at least nine diseases and disease groups listed by the World Health Organization (WHO) (8). More than 1 billion people are known to be either at risk or infected by skin NTDs (9). They mainly prevail in poor communities of low- and middle-income countries (LMICs) where resources are scarce and where there are limited numbers of dermatologists to diagnose the conditions. Additionally, skin NTDs more often affect people of color. Availability of screening systems, therefore, is critical for this set of diseases which will enable earlier diagnosis and treatment. The longer the delay in diagnosis, the more patients with skin NTDs may be left with life-long disabilities and deformities.

While there is growing interest in the use of deep learning for diagnosis of skin NTDs to fill in these gaps, there have been limited studies to date which investigated the development of an AI model for a combination of these less studied diseases in the less studied populations. In this study, we aimed to develop deep learning based AI models with clinical images we collected for five skin NTDs, namely, Buruli ulcer, leprosy, mycetoma, scabies, and yaws, to understand how diagnostic accuracy is influenced by different models, especially when the training images are relatively small in number and collected under diverse conditions. All of the images are from dark-skinned African populations, with Fitzpatrick skin type IV or above. We anticipate that our findings will support future development of AI models for the skin NTDs, and in addition, other skin diseases in people with darker skin types.

## METHODS

This study used photographs that were collected prospectively in the West African countries of Côte d’Ivoire and Ghana, through our ongoing studies with use of digital health tools for clinical data documentation and for teledermatology. The description of the design of this study can be found elsewhere (10). Briefly, photographs of skin lesions were collected with clinical information including demographics and disease description to support dermatologists in providing diagnoses remotely. The photographs were taken using the camera on Lenovo Tab M10 FHD Plus smart tablets under field conditions and in rural clinics from a total of six health districts (four in Côte d’Ivoire and two in Ghana) known to be endemic with one or more skin NTDs. Image resolution was 1920 × 2560 pixels stored in JPEG format. Written informed consent was obtained from all patients for use of their images. The study has ethical approvals from the institutional review board of the Tulane University School of Public Health and Tropical Medicine (2020-2054-SPHTM) (USA), the Ministry of Health of Côte d’Ivoire (No. IRB000111917), and the Ministry of Health of Ghana (GHS-ERC:014/05/21).

### Dataset screening

Images were selected from our data repository for which diagnoses for one of the five targeted diseases (Buruli ulcer, leprosy, mycetoma, scabies, and yaws) were made remotely and in person by two dermatologists with more than 10 years of experience in diagnosing patients locally. A portion of cases of Buruli ulcer underwent polymerase chain reaction (PCR) testing for confirmation. Likewise, dual path platform (treponemal and non-treponemal) (DPP) testing (Chembio Diagnostics, Medford, NY, USA) was done for a portion of cases of yaws. Table 1 shows the data summary of the five diseases, with number of patients and number of images for each disease. Multiple images were obtained for most patients. For Buruli ulcer and yaws, the numbers within parenthesis were those with positive results with PCR and DPP, respectively.

**Table 1.**
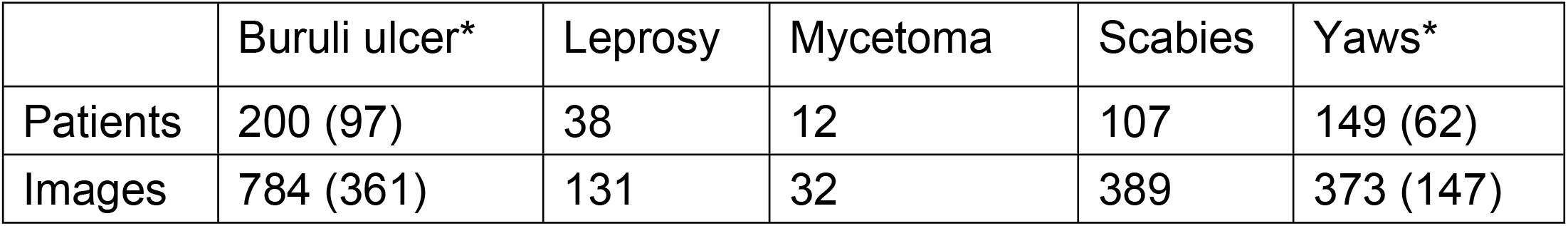
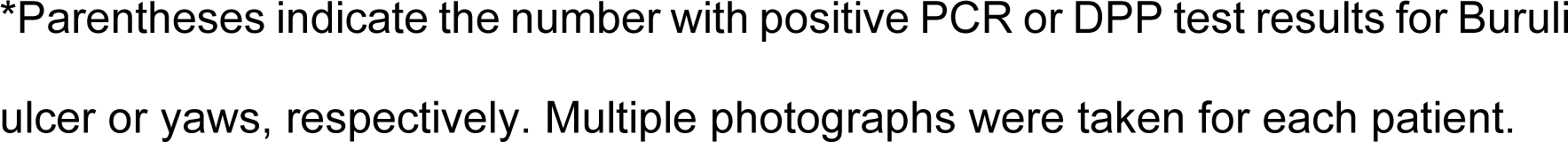
Patient and image sample sizes

|  | Buruli ulcer* | Leprosy | Mycetoma | Scabies | Yaws* |
| --- | --- | --- | --- | --- | --- |
| Patients | 200 (97) | 38 | 12 | 107 | 149 (62) |
| Images | 784 (361) | 131 | 32 | 389 | 373 (147) |

\*Parentheses indicate the number with positive PCR or DPP test results for Buruli ulcer or yaws, respectively. Multiple photographs were taken for each patient.

### AI-based skin disease diagnosis model

Convolutional neural networks (CNNs) are the popular deep learning techniques to extract feature representation from the image samples for disease diagnosis. CNNs are multi-layer neural networks with convolutional filters to capture the visual pattern from skin images. In this study, we adopted two popular CNNs, the ResNet-50 (50-layer residual neural network) (11) and the 16-layer VGG-16 model (12). The purpose was to examine the performance of different deep learning architectures and validate the feasibility of deep learning models in diagnosis of these skin diseases.

All the original images were resized into the same size as a 3D tensor with 224 × 224 × 3 pixel resolution to fit the input of deep learning models. Data augmentations and normalization pre-processing strategies were also employed following existing image classification tasks (13). These were then sent to the Resnet-50 model or VGG-16 model, pre-trained on the ImageNet dataset, which is a large-scale, open-source image repository (14). Each image was represented as a 2048-dimensional feature vector for ResNet-50, and a 4096-dimensional feature vector for VGG-16. Following this, we designed the disease diagnosis classifier with output of a 5-dimensional vector as a 5-disease probability vector. For model optimization, we adopted stochastic gradient descent (SGD) with a momentum of 0.9 as optimizer to update the whole network parameters (*i.e*., ResNet50 and classifier parameters). We performed the experiments using the PyTorch library running on one GPU (NVIDIA Titan V).

To train our designed models, the images from k% of patients from our collection chosen at random was used as a training set, and the images from the remaining patients were used to evaluate the model performance. We tested if and how laboratory confirmation may change the accuracy of the classifier. With our datasets (Table 1), we performed two kinds of experiments: firstly, using all cases [clinical diagnosis] (Task 1), and secondly, using only those cases that tested positive with PCR or DPP for Buruli ulcer and yaws, respectively [test positives] (Task 2). Otherwise, the analysis was the same. There was no patient overlap between the training and test sets.

We adopted two metrics, the Top-1 accuracy (%) and the Matthew’s correlation coefficient (MCC, 0∼1) to evaluate our model (15). Top-1 accuracy measures the proportion of test images for which the predicted disease matches the single target disease. MCC is a reliable statistical score that produces a high value only if the prediction obtained good results in all the four confusion matrix categories (true positives, false negatives, true negatives, and false positives). To map the learned visual representations, we used the dimensionality reduction method, Principal Component Analysis (PCA) (16).

## RESULTS

Table 2 presents the results of diagnostic accuracy from the two models (ResNet-50 and VGG-16) using all images as Task 1 and using images from laboratory confirmed positive cases of Buruli ulcer and yaws in training as Task 2. From the results across Tasks 1 and 2, we observed a consistent performance improvement when we had more training samples.

**Table 2:** Diagnostic accuracy of the two network models with different percentages of images for training

| k% | 30 | 40 | 50 | 60 | 70 |
| --- | --- | --- | --- | --- | --- |
| Top-1 Acc | 78.55% | 79.36% | 82.20% | 82.84% | 84.63% |
| MCC | 0.6776 | 0.6897 | 0.7308 | 0.7394 | 0.7715 |

VGG-16
| k% | 30 | 40 | 50 | 60 | 70 |
| --- | --- | --- | --- | --- | --- |
| Top-1 Acc | 76.61% | 78.45% | 80.55% | 80.27% | 82.22% |
| MCC | 0.6440 | 0.6758 | 0.7038 | 0.6984 | 0.7336 |

(b) Task 2: Lab test positives ResNet-50
| k% | 30 | 40 | 50 | 60 | 70 |
| --- | --- | --- | --- | --- | --- |
| Top-1 Acc | 75.71% | 76.52% | 77.19% | 79.15% | 84.17% |
| MCC | 0.6505 | 0.6615 | 0.6725 | 0.7011 | 0.7706 |

VGG-16
| k% | 30 | 40 | 50 | 60 | 70 |
| --- | --- | --- | --- | --- | --- |
| Top-1 Acc | 73.64% | 74.93% | 75.79%% | 78.72% | 79.44% |
| MCC | 0.6204 | 0.6407 | 0.6515 | 0.6948 | 0.7011 |

ResNet-50 and VGG-16 are two CNN image models with the former having the higher number of layers. K% is the percentage of all images used in training. Training and test samples did not overlap. Top-1 Acc: top-1 accuracy is the proportion of test images for which the predicted diagnosis by the model matches the actual diagnosis. MCC: Matthew's correlation coefficient is a statistical value assessed by results obtained from four confusion matrix categories. A higher value means better prediction (range, 0-1).

### Confirmatory analysis and confusion matrix

To further understand our AI diagnosis model for each disease, we analyzed the confusion matrices of the two models to examine if the model trained with images from the diagnostic test positive data would contribute to better performance. For Setting A, we used 50 PCR positive Buruli ulcer patients with PCR positives plus the other 4 diseases to train the model based on ResNet-50. For Setting B, we used 50 clinically diagnosed Buruli ulcer patients (including every patient diagnosed as Buruli ulcer irrespective of their PCR results) plus the other 4 diseases to train the model based on ResNet-50. The test set was the same for the two models, which included 100 clinically diagnosed Buruli ulcer patients plus the other 4 diseases. There was no patient overlap between the training and test set. Overall, Setting A was able to achieve 80% accuracy while this was 77% for Setting B.

Figure 1 shows confusion matrices by each disease. Each row of the matrix represents the instances of actual diagnosis [ground truth], while each column represents the instances of predicted diagnosis by the deep learning model [prediction]. Each diagonal element denotes the correct diagnosis by the model per disease. The prediction accuracy increased in Setting A by 1-3% as compared to Setting B across all diseases besides for mycetoma, where there were smaller number of images. For both Settings A and B, Buruli ulcer and scabies had the highest percentages of correct diagnosis, 88% and 85% for both, respectively.

**Figure 1.**
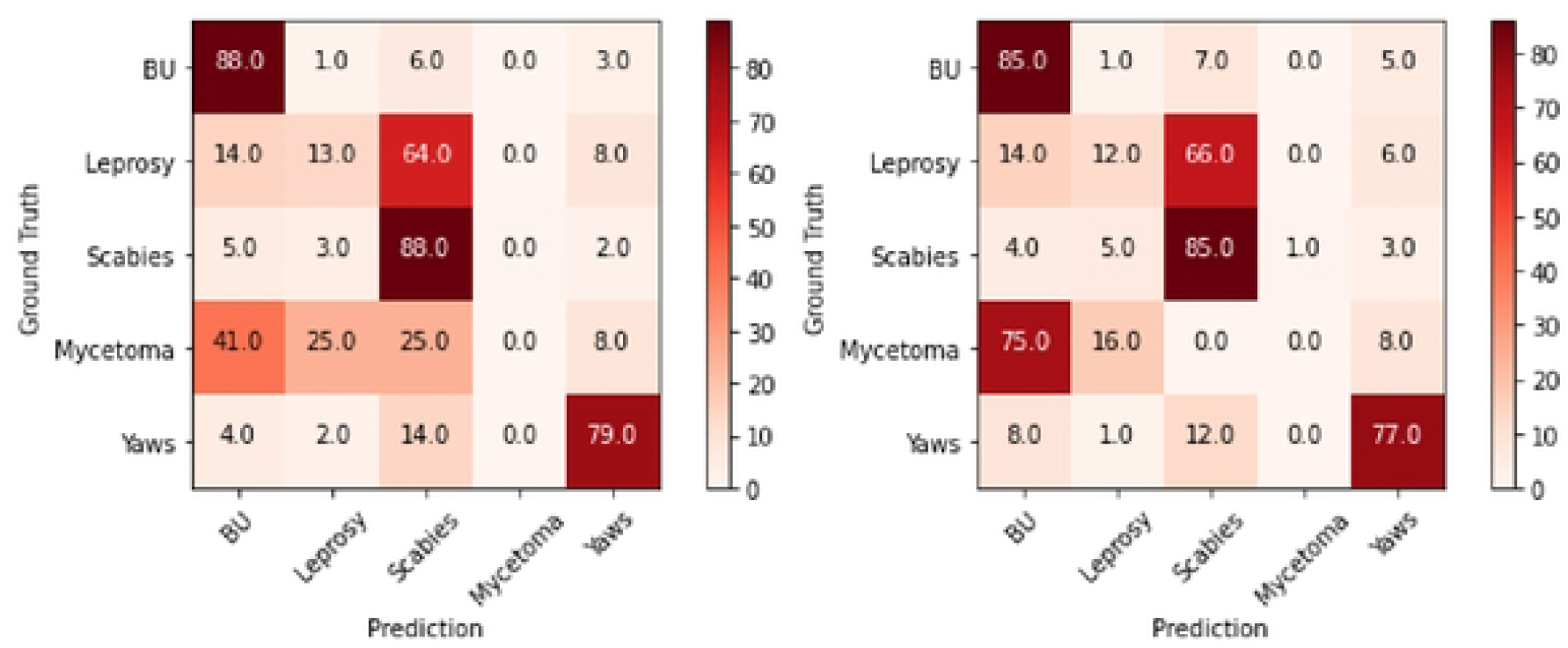
Confusion matr ces showing the proportions of diagnosis in the original images [ground truth] and the predicted diagnosis by the deep learning model [prediction]. Left: Setting A, using a model trained with PCR-positive Buruli ulcer cases. Right: Setting B, using a model trained with clinically diagnosed Buruli ulcer cases.

### Qualitative Analysis

Figure 2 provides eight example images which resulted in incorrect prediction by our pilot AI model based on ResNet-50, with (k=50)% training data for all data (Task 1). Numbers in the parentheses represent the likelihood of the diagnosis by the prediction model [prediction label] as compared to the actual diagnosis [true label], or the ground truth. An uncertainty score is also given to each test image, which is calculated by the correlation between the predicted probability with random guess. Higher correlation means higher uncertainty score. The uncertainty score indicates the degree of irrelevant evidence the AI model finds for the given test image used to predict its diagnosis. For example, Figure 2(a) shows a true label score for yaws of 0.187 and a predicted label for Buruli ulcer of 0.254 with high uncertainty of 0.93. This means that the model predicted the image to be more like Buruli ulcer than yaws, however it was also highly uncertain. An uncertainty score closer to 1 represents higher uncertainty for the diagnosis output. When it is 100% uncertain, AI estimates it to be a random guess and provides a confidence score of 0.200 (5 diseases, 1/5 = 0.200). The AI prediction is better when the uncertainty score is lower, although the diagnosis could still be incorrect.

**Figure 2.**
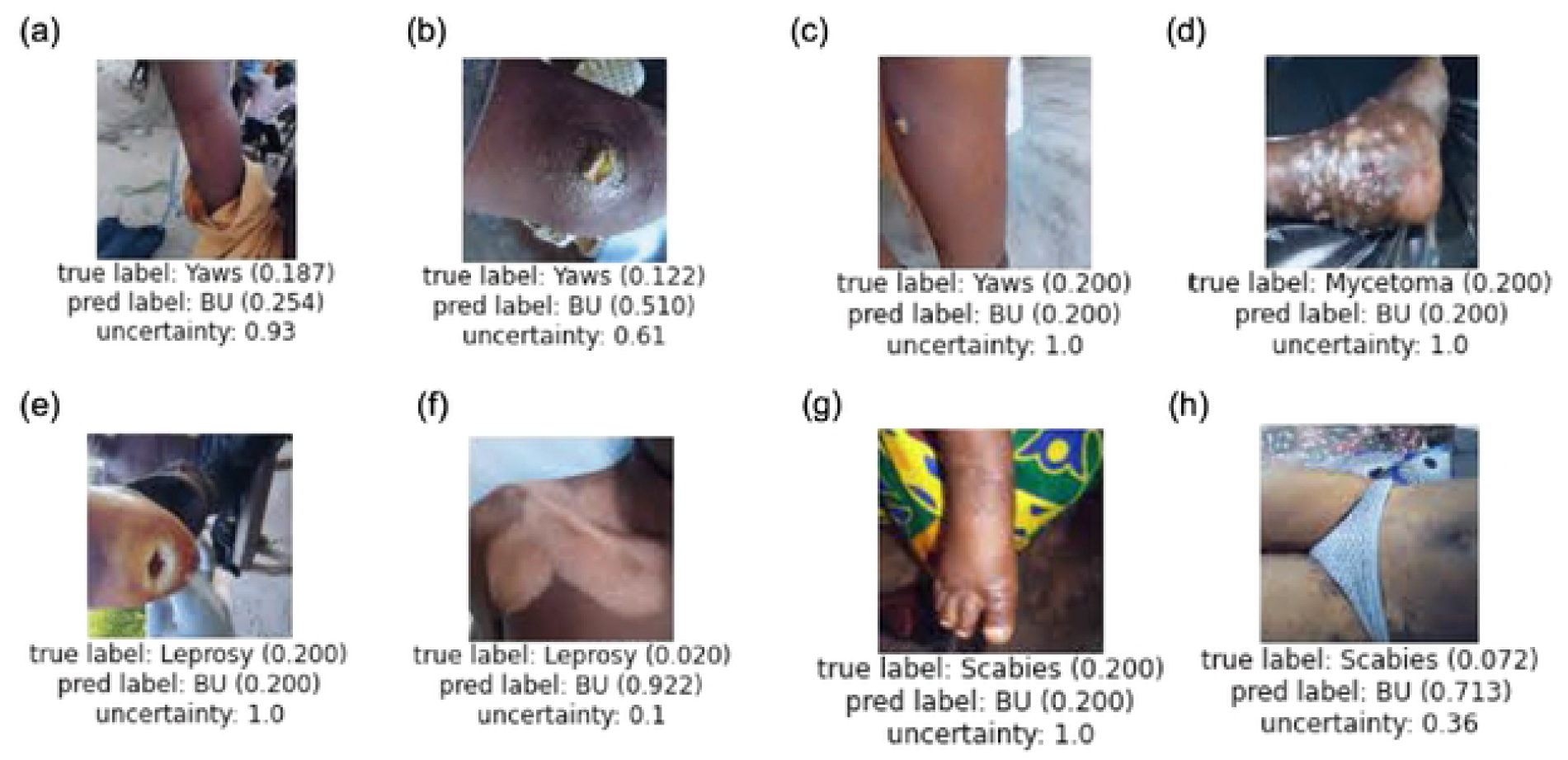
Examples of incorrect prediction made by our Al-based diagnosis model based on ResNet50, with (k=50)% training data for all data (Task 1).

### Feature Visualization

To further understand why we can achieve better performance on Buruli ulcer and scabies but worse performance for instance on mycetoma, we used, PCA (16) to map the learned visual representations (2048-dimensional features of ResNet-50) of each test class image to a 2-D plane. The goal was to visualize the learned feature representation and provide a direct way to understand the discriminative ability of AI features from raw skin images. Figure 3a shows the training samples while Figure 3b lists the test samples, where each dot in the 2-D plane denotes one image sample and the same color means the image samples of the same disease. From the results, we observed that our classification model can learn discriminative features from raw skin images to differentiate diseases in the training stage, while the model generalization ability to test images becomes poorer, which means the model cannot easily differentiate the test images like the training ones.

**Figure 3.**
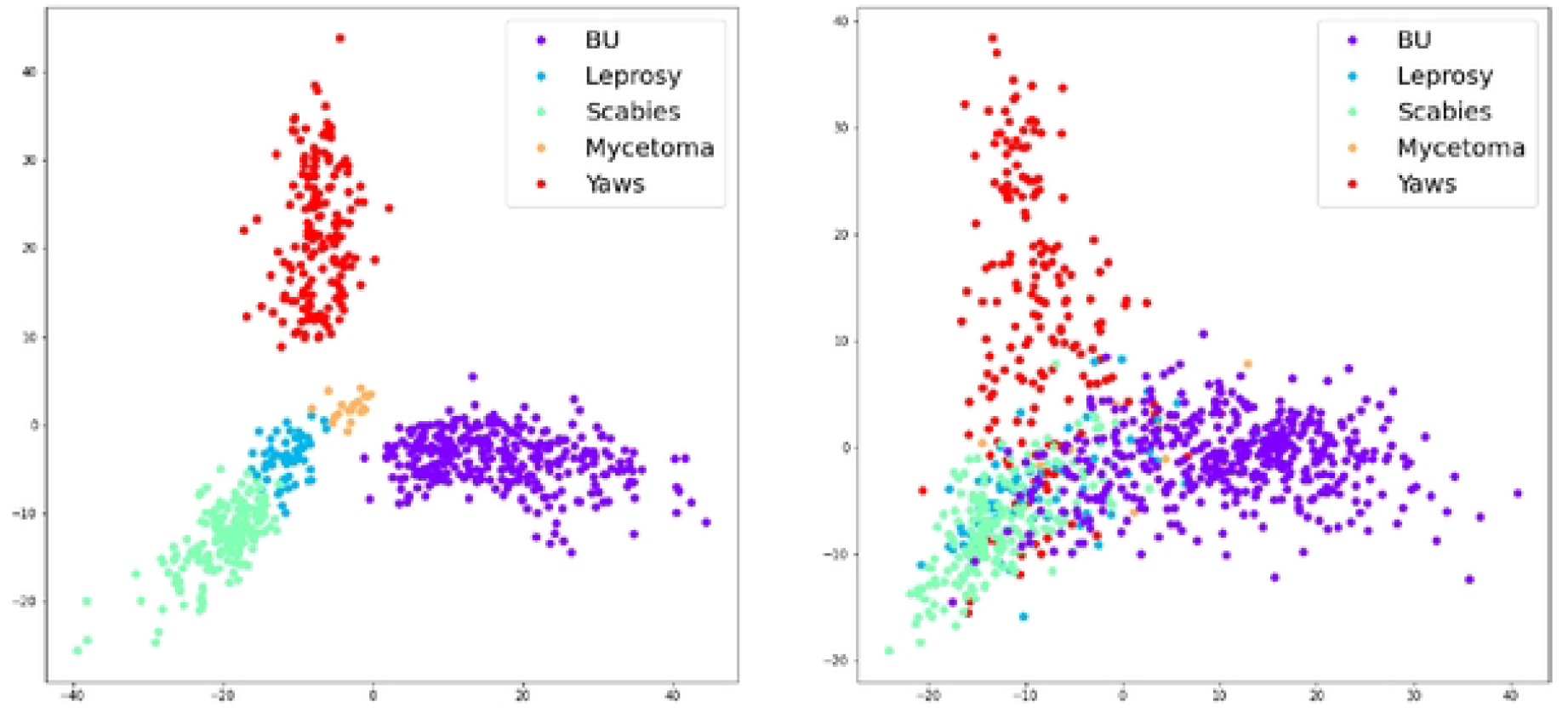
Mapping out diagnostic accuracy using Principal Component Analysis. (a)Training samples from 5 diseases. (b) Test samples from 5 diseases.

## DISCUSSION

In this report, we explored how deep learning might help in screening and/or diagnosis of skin NTDs, which often affects people with darker skin tones. Two deep learning models were examined in our work. Between the ResNet-50 and VGG-16 models, we conclude that the ResNet-50 model achieved better performance (around 2% better prediction for all evaluation) in predicting our skin images. The major difference between the two models is the depth of their layers, *i.e*., ResNet-50 contains 50 layers of convolutional, pooling operations, while VGG-16 only contains 16 layers of the same. Generally, deeper models with more layers can extract more powerful representations from image data (12). This tendency was consistent also for our dataset which focused on skin disease diagnosis. However, models with more layers contain more parameters, which make them heavier, and less efficient (12). VGG-16 is more efficient as fewer layers are included.

Although classified together as skin NTDs, the target infections have quite different appearances, presentations, and progressions. The lesions can be raised, depressed, smooth or rough, various colors or multicolored even for the same condition. We observed that deep learning approaches to identification of Buruli ulcer, scabies and yaws showed good performance of close to over 80% prediction, perhaps since these were trained with more images. Especially for Buruli ulcer where there were 784 images, the performance was over 90%. Leprosy and mycetoma, used smaller sample sizes and had poorer performance. For leprosy, we speculate that it was not only the sample size, but also the complexity of the disease presentation that impacted performance (17). We had a range of images from tuberculoid to borderline to lepromatous type leprosy, as well as some included deformities and wounds that developed due to peripheral neuropathy. We stratified these different conditions and ran the same analysis, with an expectation that this may increase power by reducing variance. However, this further decreased the number of our samples, and we were unable to obtain any meaningful results this time. Likewise, similar results were obtained for yaws, when stratified for ulcerative versus non-ulcerative (papilloma, hyperkeratosis, etc.) lesions. However, we believe if there were enough images, stratifying may increase the accuracy of the predicted diagnosis. Moreover, as prior study on AI-based diagnosis for leprosy showed, clinical data other than images, most importantly loss of sensation for leprosy, are essential to be combined in the deep learning dataset for better model development (17).

The percentages of correct diagnosis increased with PCR-positive cases of Buruli ulcer. This demonstrated that it may be better to input images from the more accurately diagnosed cases in the training models also for achieving better accuracy in the generated AI models. It was interesting to see that the PCR-confirmed cases of Buruli ulcer contributed in increasing the diagnostic accuracy not just for Buruli ulcer but also for other diseases. On the other hand, contrary to our hypothesis, the percentage increase was minimal (3% for Buruli ulcer), which may be an indication that the accuracy of clinical diagnosis alone is reliable to an extent. Especially for Buruli ulcer, a previous study by Eddyani et al. has shown that sensitivity of clinical diagnosis was as high as 92% (95% CI, 85-96%), which was the highest among any other methods including PCR (18). PCR results can be false negatives in Buruli ulcer due to several factors, for example, site of sample collection, skills in sample taking and duration of the wound (19). While it is currently the preferred test for diagnostic confirmation, it has its flaws and is not always reliable. In many studies, PCR is considered 65-70% sensitive (20) or even only 61% sensitive (21). Specificity is perhaps highest for the PCR positive cases, but sensitivity is highest for clinically identified cases. The PCR positive cases should be enriched for true cases, but it also misses true cases. One hope for AI – which our findings also support – is that it will objectively resolve this gap between diagnostic tests like PCR and clinical diagnoses with addition of another tool.

Incorrect diagnoses made by our model were skewed towards other skin NTDs being diagnosed as Buruli ulcer, as about half of our images were Buruli ulcer. Fairness issues in deep learning arise when the dataset is extremely imbalanced across different categories or groups (22). When these images with incorrect prediction were reviewed, some cases would have been difficult to differentiate even with the human eye, such as the case of yaws in Figure 2(b), for example. On the other hand, some cases with obviously different presentations were predicted to be Buruli ulcer, such as the case of mycetoma in (d) and leprosy in (f). We were unsure why they were predicted to be Buruli ulcer.

For cases shown in (a) and (g), images and location on the limbs may have played some role in these being predicted as a Buruli ulcer case, as the most commonly affect body parts in Buruli ulcer are the limbs (23, 24). Figure 2(c) was a case of yaws, but the main lesion was not centered, and the lesion of interest was not very obvious. The backgrounds or the clothes may have disturbed the predictions in cases such as in (a), (e), (g), and (h). It will be necessary to understand these patterns in order to resolve incorrect predictions, which will be one of our future study directions.

A major source of bias in AI applications stems from the availability and variety of images used in training. There are a very limited number of images of these diseases and a more limited number of images of people of color. In addition, the phrase “people of color” embraces a huge range of hues and surface characteristics even within the African continent. One of the strong points of this pilot has been the use of local dermatologists. In one example in the field, the local dermatologists recognized a series of deeply pigmented lesions as being a reaction to skin whitening agents, a diagnosis that would not easily be arrived at by physicians in the US or Europe. A key observation here has been to reinforce the need for more images from a wider diversity of cases from this part of the world, similarly to the recently recognized gap in dermatological training in general (25, 26). We were able to derive almost the same, if not close, accuracy in diagnosis with the model trained with images from clinical diagnosis over those trained with images with laboratory confirmation – this was partly possible because of the involvement of our skilled local dermatologists.

There are limitations to our study, some of which were already described, such as limited number of images and imbalance in image numbers between diseases. Moreover, images were taken under different conditions, and they were highly heterogeneous, for example, distracting objects in the background or lighting. We are currently working on how to mitigate them, as the photos are taken under field conditions in Côte d’Ivoire and Ghana where conditions are less formal than with many other studies. As it is difficult to mandate the images be taken in a uniform environment in these settings, and as this will also limit the number of images that we can use for deep learning, it is potentially more up to the technology how we can overcome this challenge. If such technology can be developed, it could be beneficial for the development of deep learning models for a wide range of skin diseases that are common in the developing world, in addition to what were targeted in this study.

## Conclusions

Here, we presented our exploratory approach in developing deep learning models for skin NTDs and the challenges that we encountered. These attempts have only just begun. We hope that the lessons learnt here will support the future development of AI technology for these neglected diseases in the neglected populations. Our approach was to have the deep learning model distinguish between multiple pathologies simultaneously. This is different from many other studies where deep learning models were asked to make a diagnosis of a single disease. However, in real-world, what happens in clinicians’ mind is that we are required to compare between different pathologies – accordingly, we devised an approach that is more in line with this practice. AI is not yet a replacement for human diagnosis, but if used well and appropriately, it is a tool that can be useful in screening for diseases and improving patient outcomes.

Particularly, the hope is that it will address the unmet needs where access to medical care is limited, like for those affected by skin NTDs.

## Data Availability

The data sets generated or analyzed during this study are available from the corresponding author on reasonable request.

## Acknowledgements

We would like to pay special thanks to Prof. Bamba Vagamon and Prof. Almamy Diabate (Université Alassane Ouattara, Service de Dermatologie CHU de Bouaké-Côte d’Ivoire) for their support in diagnosis of skin diseases on site and remotely. We would like to also thank the project team, Aubin Yao and Luc Kowaci Gontran Yeboue (Hope Commission International) for their hard work in implementing the teledermatolgy project.

## References

1. Patel S, Wang JV, Motaparthi K, Lee JB. Artifical Intelligence in dermatology for the clinicians. Clinics in Dermatology. 2021;39:667–72.

2. Esteva A, Kuprel B, Novoa RA, Ko J, Swetter SM, Blau HM, et al. Dermatologist-level classification of skin cancer with deep neural networks. Nature. 2017;542(7639):115–8.

3. Haenssle HA, Fink C, Schneiderbauer R, Toberer F, Buhl T, Blum A, et al. Man against machine: diagnostic performance of a deep learning convolutional neural network for dermoscopic melanoma recognition in comparison to 58 dermatologists. Ann Oncol. 2018;29(8):1836–42.

4. Brinker TJ, Hekler A, Enk AH, Berking C, Haferkamp S, Hauschild A, et al. Deep neural networks are superior to dermatologists in melanoma image classification. Eur J Cancer. 2019;119:11–7.

5. Jones OT, Matin RN, van der Schaar M, Prathivadi Bhayankaram K, Ranmuthu CKI, Islam MS, et al. Artificial intelligence and machine learning algorithms for early detection of skin cancer in community and primary care settings: a systematic review. Lancet Digit Health. 2022;4(6):e466–e76.

6. Dick V, Sinz C, Mittlbock M, Kittler H, Tschandl P. Accuracy of Computer-Aided Diagnosis of Melanoma: A Meta-analysis. JAMA Dermatol. 2019;155(11):1291–9.

7. Seite S, Khammari A, Benzaquen M, Moyal D, Dreno B. Development and accuracy of an artificial intelligence algorithm for acne grading from smartphone photographs. Exp Dermatol. 2019;28(11):1252–7.

8. World Health Organization. Ending the neglect to attain the Sustainable Development Goals: a strategic framework for integrated control and management of skin-related neglected tropical diseases. Geneva, Switzerland; 2022. Contract No.: Licence: CC BY-NC-SA 3.0 IGO.

9. Yotsu R, Fuller LC, Murdoch ME, Revankar C, Barogui Y, Pemmaraju VRR, et al. WHO strategic framework for integrated control and management of skin-related neglected tropical diseases (skin NTDs). What does this mean for dermatologists? British Journal of Dermatology. 2023; 188(2): 157–159.

10. Yotsu RR, Itoh S, Yao KA, Kouadio K, Ugai K, Koffi YD, et al. The Early Detection and Case Management of Skin Diseases With an mHealth App (eSkinHealth): Protocol for a Mixed Methods Pilot Study in Cote d’Ivoire. JMIR Res Protoc. 2022;11(9):e39867.

11. He K, Zhang X, Ren S, Sun J, editors. Deep residual learning for image recognition. IEEE Conference on Computer Vision and Pattern Recognition; 2016; Las Vegas, NV, USA.

12. Simonyan K, Zisserman A. Very Deep Convolutoinal Networks for Large-scale Image Recognition. arXiv preprint. 2015(1409):1556.

13. Ding Z, Liu H, editors. Marginalized Latent Sematic Encoder for Zero-Shot Learning. IEEE/CVF Conference on Computer Vision and Pattern Recognition; 2019; Long Beach, CA, USA.

14. Deng J, Dong W, Socher R, Li LJ, Li K, Fei-Fei L, editors. ImageNet: A large-scale hierarchical image database. IEEE Comput Vis Pattern Recognit; 2009; Miami, Florida, USA.

15. Chicco D, Jurman G. The advantages of the Matthews correlation coefficient (MCC) over F1 score and accuracy in binary classification evaluation. BMC Genomics. 2020;21(1):6.

16. Abdi H, Williams LJ. Principal component analysis. Wires Computational Statistics. 2010;2(4):433–59.

17. Barbieri RR, Xu Y, Setian L, Souza-Santos PT, Trivedi A, Cristofono J, et al. Reimagining leprosy elimination with AI analysis of a combination of skin lesion images with demographic and clinical data. Lancet Reg Health Am. 2022;9:100192.

18. Eddyani M, Sopoh GE, Ayelo G, Brun LVC, Roux JJ, Barogui Y, et al. Diagnostic Accuracy of Clinical and Microbiological Signs in Patients With Skin Lesions Resembling Buruli Ulcer in an Endemic Region. Clinical infectious diseases : an official publication of the Infectious Diseases Society of America. 2018;67(6):827–34.

19. van der Werf TS. Diagnostic Tests for Buruli Ulcer: Clinical Judgment Revisited. Clinical infectious diseases : an official publication of the Infectious Diseases Society of America. 2018;67(6):835–6.

20. Bretzel G, Siegmund V, Nitschke J, Herbinger KH, Thompson W, Klutse E, et al. A stepwise approach to the laboratory diagnosis of Buruli ulcer disease. Trop Med Int Health. 2007;12(1):89–96.

21. Siegmund V, Adjei O, Nitschke J, Thompson W, Klutse E, Herbinger KH, et al. Dry reagent-based polymerase chain reaction compared with other laboratory methods available for the diagnosis of Buruli ulcer disease. Clinical infectious diseases : an official publication of the Infectious Diseases Society of America. 2007;45(1):68–75.

22. Jing T, Xu B, Li J, Ding Z. Towards Fair Konwledge Transfer for Imbalanced Domain Adaptation. IEEE Transactions on Image Processing. 2021;30:8200–11.

23. Hospers IC, Wiersma IC, Dijkstra PU, Stienstra Y, Etuaful S, Ampadu EO, et al. Distribution of Buruli ulcer lesions over body surface area in a large case series in Ghana: uncovering clues for mode of transmission. Trans R Soc Trop Med Hyg. 2005;99(3):196–201.

24. Sexton-Oates NK, Stewardson AJ, Yerramilli A, Johnson PDR. Does skin surface temperature variation account for Buruli ulcer lesion distribution? PLoS Negl Trop Dis. 2020;14(4):e0007732.

25. Chang MJ, Lipner SR. Analysis of Skin Color on the American Academy of Dermatology Public Education Website. Journal of drugs in dermatology : JDD. 2020;19(12):1236–7.

26. Okoji UK, Lipoff JB. Demographics of US dermatology residents interested in skin of color: An analysis of website profiles. J Am Acad Dermatol. 2021;85(3):786–8.

